# Estimating the impact of a denicotinisation policy on smoking behaviours in Australia – an expert knowledge elicitation study

**DOI:** 10.64898/2026.07.21.26358615

**Authors:** Samantha Howe, Richard Edwards, Kylie Morphett, Cheneal Puljević, Tim Wilson, Coral Gartner, Tony Blakely, Driss Ait Ouakrim

## Abstract

**Background:** Mandatory reductions in tobacco product nicotine content to non-addictive levels may help achieve a tobacco endgame. Evidence exists for potential positive impacts of ‘very low nicotine content’ (VLNC) cigarettes in promoting smoking cessation in trial settings, but the population-level policy impact is unknown.

**Methods:** An online expert knowledge elicitation survey was undertaken to generate estimates for the impact of a VLNC mandate in Australia. Global experts in tobacco control were recruited to individually estimate impacts on smoking initiation, cessation, and switching to e-cigarettes or illicit market cigarettes. Linear pooling was used to estimate a combined (non-parametric) distribution per question, incorporating minimum, maximum, and best guess values from each respondent.

**Results:** The survey was completed by 15 experts. Interquartile ranges from pooled responses indicated that in the first year of a VLNC policy, smoking cessation could increase by 1.2-7.1 times the rate without the policy and switching to vaping by 1.3-8 times. Switching to illicitly supplied ‘normal’ nicotine cigarettes was estimated between 15-50% of those who smoke. Smoking initiation estimates varied greatly. Experts from Australia (n=8) consistently reported lower estimates for positive outcomes, and higher illicit market switching estimates, than experts from other countries.

**Conclusions:** Given the absence of implementation evidence, this study provides Australia-specific estimates of the potential behavioural effects of a VLNC policy. The findings suggest that denicotinisation would accelerate declines in smoking, but that it may be undermined by illicit cigarette access in Australia. Estimates should be updated with real-world evidence as it becomes available.

**Implications:**

- This study quantifies a range of behavioural responses to a national-level VLNC mandate in Australia, based on the knowledge of 15 global experts in tobacco control.
- The pooled responses indicate that a VLNC mandate could increase smoking cessation and switching to e-cigarette use, with variable impacts on youth smoking and vaping uptake.
- Denicotinisation could accelerate progress towards a tobacco endgame in Australia, but its benefits may be limited if illicit access to nicotine-containing cigarettes continues. Future modelling in Australia based on this analysis must explicitly consider the wide variation in expert responses, and other sources of evidence.

## Introduction

As countries pursue smokefree societies or a ‘tobacco endgame’, several novel interventions have been proposed to accelerate reductions in tobacco smoking. One such intervention is a mandatory reduction in nicotine content in cigarettes and other tobacco products to non-addictive levels, so that only ‘denicotinised’ or ‘very low nicotine content’ (VLNC) cigarettes can be sold. By reducing the addictiveness and appeal of tobacco products, a VLNC mandate aims to increase cessation and reduce smoking uptake.

The Australian Government has included consideration of a nicotine reduction policy in the National Tobacco Strategy 2023-2030 (Priority Area 7.6 [1]) but has not committed to implementation. Aotearoa/New Zealand was the first country to legislate a VLNC standard in 2022, however the legislation was repealed before implementation.[2] The United States Food and Drug Administration (FDA) issued a proposed denicotinisation rule in January 2025, with an intended implementation date in 2027 [3], but this has been indefinitely paused. No jurisdiction has yet successfully implemented a mandatory VLNC standard.

Randomised controlled trials (RCTs) indicate that VLNC cigarettes can promote smoking cessation and switching to alternative products including e-cigarettes.[4,5] However, RCTs cannot directly estimate the population-level effects of a mandatory VLNC standard. Trial conditions may poorly reflect policy implementation. VLNC cigarettes are often supplied free of charge, and participants can usually obtain ‘normal’ nicotine content cigarettes, with non-adherence being common (e.g. [6, 7]). Most trials are short, limiting inference about long-term behaviour. Recruitment criteria may produce samples that differ from the broader population who smoke, including studies restricted to people not intending to quit.[8] Finally, it is not possible to estimate the impact of VLNC cigarettes on smoking uptake in a trial setting.

Expert Knowledge Elicitation (EKE, also known as structured expert elicitation) can quantify expected policy effects when primary evidence or natural experiments are unavailable or insufficient. [9] Tobacco control EKEs have previously estimated the population-level effect of the FDA’s proposed VLNC standard [10,11], and menthol ban.[12] EKE also allows context-specific factors to be incorporated. For Australia, two factors are particularly important. First, the potential undermining effect of illicitly traded tobacco, which has increased considerably over the past decade [13] and was recently estimated to account for 50-60% of total tobacco consumption in 2024-25.[14] Second, the availability of e-cigarettes as a substitute product within a legal supply model restricted to pharmacies (though in reality primarily accessed via the illicit market).[15,16]

This study aimed to estimate the potential effects of a VLNC policy on smoking and nicotine-use behaviours in Australia, drawing on experts in tobacco and e-cigarette research and policy, addiction, and illicit markets. Specifically, we elicited expected impacts on smoking cessation and uptake, e-cigarette use, and illicit tobacco sourcing.

## Methods

This EKE was administered via an online survey. We followed the UK Medical Research Council protocol developed by Bojke et al. [17] and drew on previous tobacco control EKEs [10,12], and guidance from the International Society for Pharmacoeconomics and Outcomes Research and the University of York on structured elicitation for healthcare decision making.[18,19] The study was designed to elicit effect sizes for a VLNC policy intervention model using SHINE-Tobacco, an existing tobacco policy simulation platform.[20,21]

### Recruitment

Recruitment occurred from August-September 2025. Eligible participants had expertise in at least one of the following areas: tobacco control research and policy, illicit tobacco markets, or nicotine addiction. We used purposive sampling, primarily based on Scopus citations (search terms in Supplementary Methods Table 2) supplemented by a rapid literature review conducted during study conceptualisation (see Supplementary Methods) and the investigators’ networks.

Experts from any country were eligible. To ensure adequate Australian contextual knowledge, the Scopus search was initially limited to Australian tobacco control experts – this group was treated as 50% of the recruitment population. There are no fixed sample-size recommendations for individual elicitation with mathematical aggregation. We aimed to recruit 30 experts, with a minimum of 15. Email invitation, including the plain language statement and consent form, were sent to 47 experts. One reminder was sent after two weeks.

Experts who agreed to participate received an individual survey link and training materials (Supplementary Material). These materials summarised the Australian policy context (based on information available at the time of recruitment), key VLNC evidence (including RCT evidence and the FDA EKE [10,22]), and instructions for completing the survey. Participants could consult additional sources if desired.

### Data collection

Data collection occurred from September 2025-March 2026 using Qualtrics XM (Version 2025). Survey questions were developed by SH, DAO and TW, then piloted by the remaining investigators (TB, CG, RE, KM, and CP), who have expertise across tobacco control policy and research, e-cigarettes, nicotine addiction, and illicit markets. Pilot testers provided feedback on the questionnaire and training materials. They were not included as participants. The final questionnaire is provided as Supplementary Material.

The survey first captured participants’ topic and country expertise and their views on whether a tobacco endgame (≤5% daily smoking prevalence) would be achieved in their country of residence within 10 years under current policies, and whether novel responses, rather than strengthened FCTC/MPOWER policies, were needed. This information was elicited to see if responses to the subsequent questions varied across these characteristics.

Participants were then asked to consider a policy in which “VLNC cigarettes, as opposed to ‘normal’ nicotine content cigarettes, are the only commercially available tobacco product on the (legal) market in Australia”. Four behavioural domains were elicited:

1. Cessation of daily smoking and dual use (i.e., concurrent smoking and vaping).
2. Switching from smoking to dual use, or from smoking to exclusive vaping.
3. Switching from legal to illicit tobacco sourcing among people who smoked or were dual users, to access normal-nicotine cigarettes.
4. Youth uptake of smoking and vaping.

Responses were elicited as event probabilities (0-100%) or relative risks compared to business-as-usual (BAU). Effect sizes were estimated separately for people <30 and 30+ years, and for the first year of fully implementation subsequent years. For some questions, BAU transition values were provided as reference points. Response summaries after each question and section allowed experts to revise their answers (examples in the Supplementary Methods).

We used the variable interval method to elicit uncertainty.[17] Experts first provided lower and upper bounds, assumed to represent 2.5^th^ and 97.5^th^ percentiles, then a ‘best guess’ assumed to represent the median. Responses were elicited individually to avoid influence from other experts. The survey was expected to take 1.5-2 hours, including reading of training materials.

### Statistical analysis

Data analysis was conducted in R (version 4.2.2).[23] Individual expert responses were plotted for each question using the SHELF package [24], to identify the most appropriate distribution for individual questions. Mathematical aggregation was then used to estimate a pooled distribution for each question, using the *qlinearpool* function (SHELF) [24] with equal weighting by expert. [17] Percentile values were reported for each question’s pooled distribution. It is important to note that the pooled distribution is not itself a parametric distribution; for example, if a beta distribution is selected for the individual responses to a given question, the pooled distribution is not a beta distribution, but instead the average of the individual beta distributions.

As a sensitivity analysis, results were aggregated separately for those who indicated high vs. low use of the training material provided alongside the survey. A post-hoc sensitivity analysis also involved excluding outliers identified for each question before pooling.

### Ethics & registration

This project was approved by the University of Melbourne’s Human Research Ethics Committee (reference number: 2025-32045-69755-3) and registered on Open Science Framework in August 2025, prior to recruitment (https://osf.io/mndte/overview).

## Results

### Participant characteristics

A total of 19 experts agreed to participate and 15 completed the survey, including eight with Australian expertise. Participants reported expertise across tobacco control, nicotine addiction, illicit markets, health economics and vaping control (Table 1).

**Table 1.**
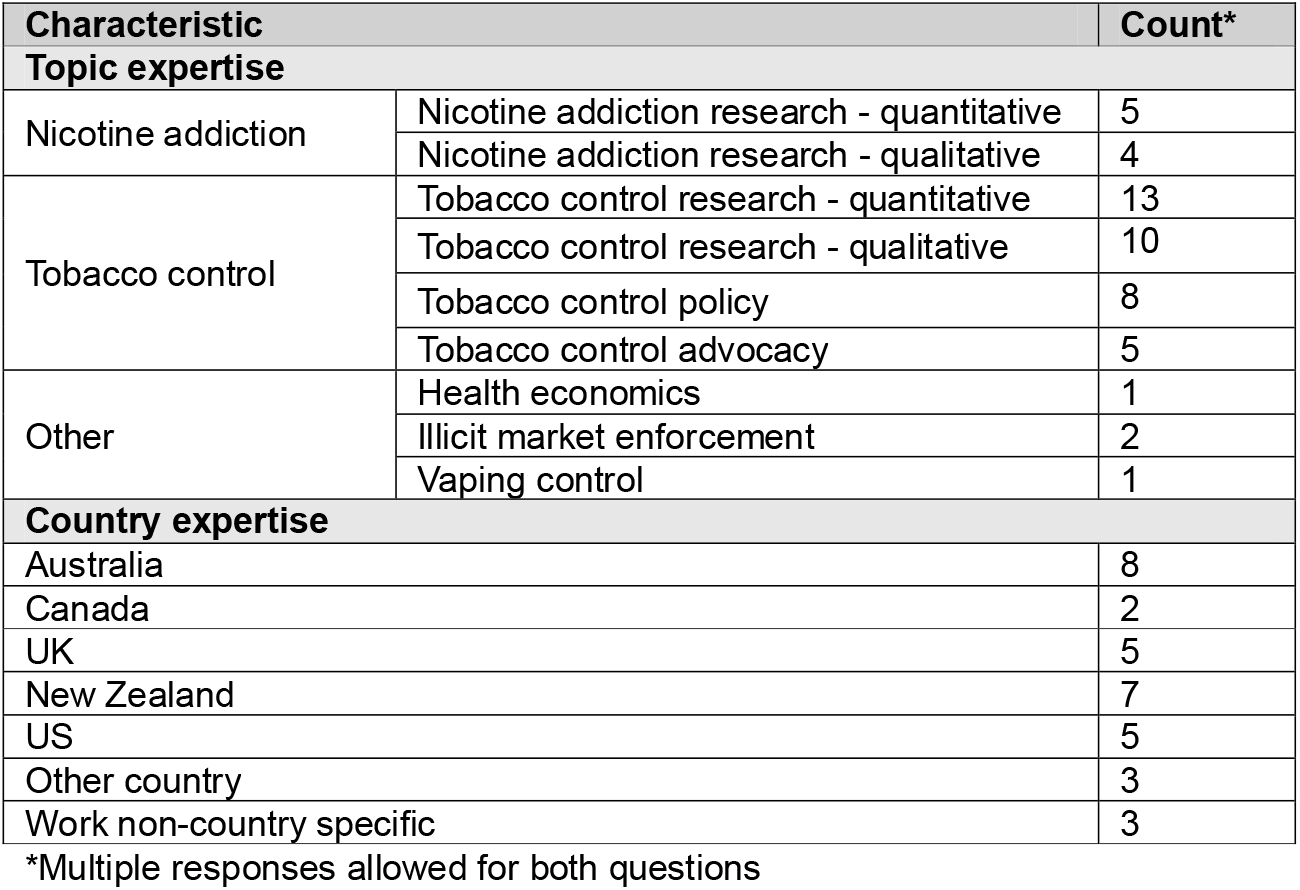
Expert characteristics.

### Belief about tobacco endgame achievability

Eleven out of 15 experts, including seven of eight Australian experts, did not expect 5% daily smoking prevalence to be achieved in their country of residence within 10 years under current policies. Twelve experts, including seven Australian experts, indicated that novel endgame policies would be required to achieve this goal.

### Impact of a VLNC policy on cessation behaviour

Table 2 presents pooled median relative risks and interquartile ranges (IQRs). For people aged 30+ years, experts estimated that a VLNC policy would increase smoking cessation to 10.2% in the first year (or twice the BAU rate, IQR 1.2-4.4) and 1.8 times BAU in subsequent years (IQR 1.1-3.1). For those aged <30 years, corresponding estimates were a 7.6% quit rate in the first year (3.6 times the BAU rate, IQR 1.7-7.1) followed by 2.9 times BAU in subsequent years (IQR 1.5-4.4). Estimated effects on cessation among dual users were smaller than effects among people who exclusively smoked (Table 2).

**Table 2.** Pooled results: median impact (and interquartile range) on smoking and vaping behaviours (n=15)

| Transition | <30-year-olds |  | 30+ year olds |  |
| --- | --- | --- | --- | --- |
|  | First year of policy | Subsequent years | First year of policy | Subsequent years |
| **Relative change compared to business-as-usual** |  |  |  |  |
| **Smoking → Former Smoking** | 3.6 (1.7-7.1) | 2.9 (1.5-4.4) | 2 (1.2-4.4) | 1.8 (1.1-3.1) |
| **Dual use → Former Smoking** | 1.9 (1.1-3.3) | 2 (1-3.7) | 1.4 (1-3.4) | 1.4 (1-3) |
| **Smoking → Vaping** | 3.3 (1.3-8) | 2.8 (1.2-5.2) | 3.8 (1.5-5.8) | 2.4 (1.2-4.8) |
| **Smoking → Dual use** | 3.8 (2.2-24.8) | 2.8 (1.2-24.3) | 3.8 (2-6.2) | 2.2 (1.2-6) |
| **No product use → Smoking\*** | 0.73 (0.02-1.0) | 0.79 (0.50-1.0) | N/A | N/A |
| **Percentage change compared to business-as-usual** |  |  |  |  |
| **Legal smoking → Illicit market<sup>#</sup>** | 25.4% (14.7%-49.8%) | N/A | 33.8% (16.3%-49.1%) | N/A |
| **Legal dual use → Illicit market<sup>#</sup>** | 22.6% (14.2%-49.8%) | N/A | 22% (8.6%-37.4%) | N/A |

\*Relative impact on smoking uptake only considered for <30-year-olds (ages for which majority of uptake occurs).
<sup>#</sup>Note that, while in Australia e-cigarettes are banned from sale in retail settings (except for pharmacy access for smoking cessation), In this study the illicit market refers to the tobacco illicit market specifically. Relative risk values <1 indicate lower transition rates under the intervention compared to BAU, while values >1 indicate a higher transition rate under the intervention compared to BAU. Interquartile range estimated by linear pooling of individual response distributions with the SHELF package in R; individual responses assumed to follow log-normal distributions, where best guess = 50<sup>th</sup> percentile, and the minimum and maximum responses are the 2.5<sup>th</sup> and 97.5<sup>th</sup> percentiles.

### Impact of a VLNC policy on switching behaviour

Estimated relative increases in switching were larger than estimated increases in quitting all nicotine (Table 2). In the first year, switching from smoking to exclusive vaping was estimated at 3.8 times BAU for people aged 30+ years (IQR 1.5-5.8) and 3.3 times BAU for those aged <30 years (IQR 1.3-8.0). Switching from smoking to dual use (i.e. taking up vaping while continuing to smoke VLNC cigarettes) was estimated at 3.8 (IQR 2.0-6.2) and 3.8 (IQR 2.2-24.8) times the BAU rates for 30+ year olds and <30-year-olds, respectively.

### Impact of a VLNC policy on illicit market sourcing

Experts estimated substantial switching to illicitly sourced normal-nicotine cigarettes. The median estimated transition to illicit sourcing in the first year of the policy was 33.8% for people aged 30+ year (IQR 16.3-49.1%), and 25.4% for those aged <30 year (IQR 14.7-49.8%). Estimates were slightly lower among dual users, but uncertainty was wide (Table 2).

### Impact of a VLNC policy on uptake

For people aged <30 years, the pooled results indicated a 27% reduction in smoking uptake in the first year compared to BAU (RR=0.73, IQR 0.02-1), followed by a 21% reduction in subsequent years (RR=0.79, IQR 0.50-1.0). Nine experts answered the follow up question on whether some young people deterred from smoking initiation due to the VLNC policy would instead take up vaping; eight answered “Yes”. The pooled median estimate was that 9.2% (IQR 1.0-19.8%) of those deterred from daily smoking due to the policy would take up vaping instead. Excluding three logically inconsistent responses from experts who had not estimated any reduction in smoking uptake reduced this estimate to 3.0% (IQR <0.01-21.7%).

### Sub-group differences

Australian experts gave more conservative estimates for quitting and switching, with all relative effect estimates being closer to the null than estimates from other country experts (Table 3). In contrast, Australian experts’ estimates for illicit market switching were approximately three times higher for people who smoked and 2.6-4.5 times higher for dual users than estimates from other country experts.

**Table 3.**
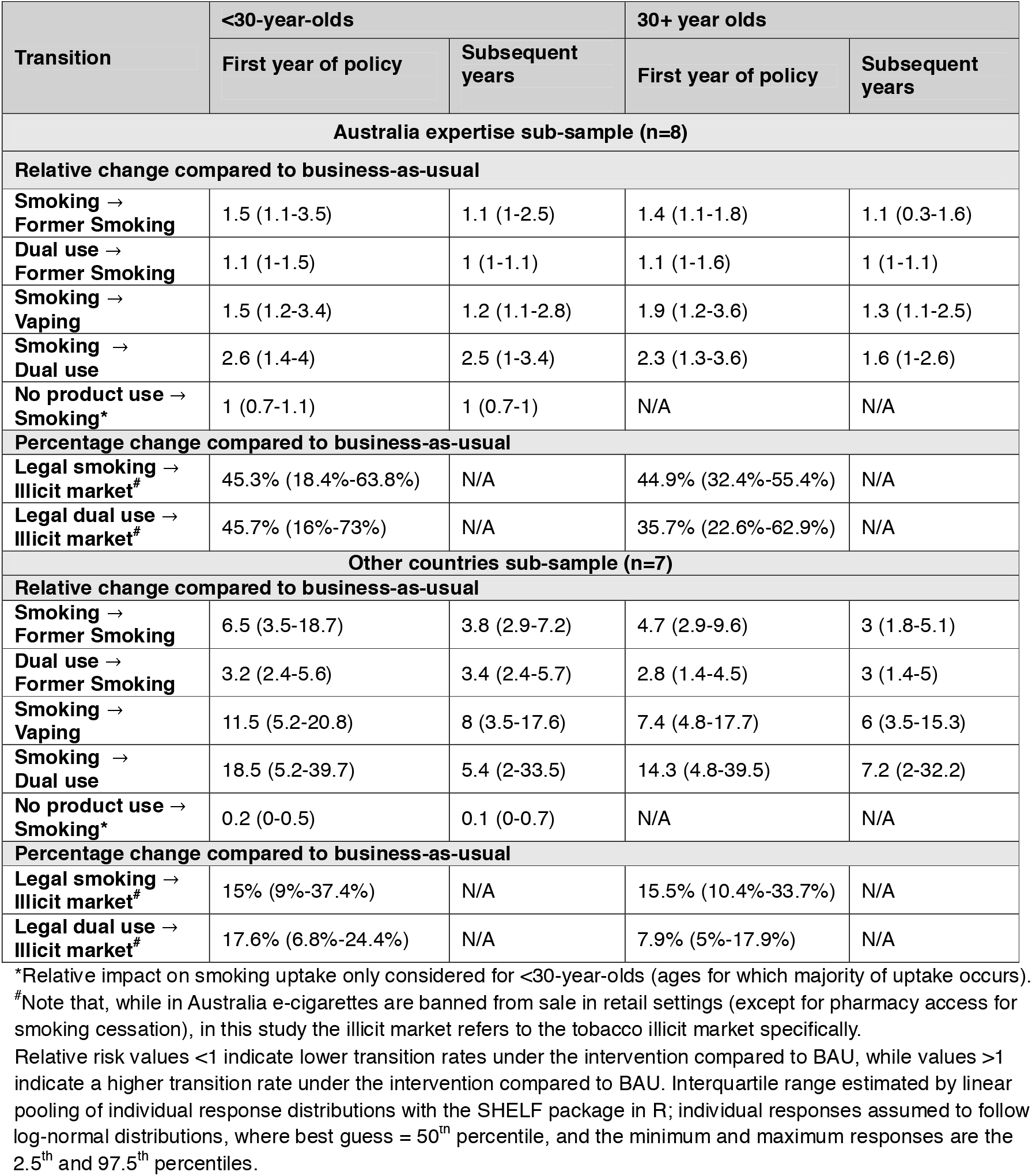
Pooled results: median impact (and interquartile range) on smoking and vaping behaviours by country expertise.

Australian experts were also more conservative about uptake. The median estimate indicated no reduction in smoking uptake in either the first or subsequent years. Comparatively, other country experts estimated a median pooled reduction by 80% in the first year of the policy and 90% in subsequent years, relative to BAU.

Restricting analyses to experts with quantitative research expertise (n=12), whom may have been more familiar with making probabilistic judgements, produced larger pooled policy effect sizes for quitting and switching than the full sample size estimates (Supplementary Table 4). The largest differences were for switching from smoking to vaping or dual use, where the median estimates were up to 1.5 times the full sample estimates, and illicit market switching for people aged <30 years, which was 1.4 times the full sample estimate (Supplementary Table 4). Uptake estimates changed little.

Finally, restricting to experts who stated that novel endgame measures were needed generally produced more conservative estimates than the main analysis (Supplementary Table 5).

### Expert feedback

Experts indicated moderate use of the training material to complete the survey (with only 1 respondent not referring to it at all). Eight of the experts did not conduct external independent research when completing the survey (Table 4). When asked how confident they felt about completing the survey, five out of 15 participants selected “Somewhat unsure – I had difficulty answering several questions”. No participants reported being very confident, and only four reported being “somewhat confident” competing the survey (Table 4).

**Table 4.** Expert survey feedback.

| Question | Response counts |  |  |  |  |  |
| --- | --- | --- | --- | --- | --- | --- |
|  | 0 = no use | 1 | 2 | 3 | 4 | 5 = high use |
| **Feedback part 1** |  |  |  |  |  |  |
| **Use of training material** | 1 | 4 | 4 | 4 | 1 | 1 |
| **Use of external research** | 8 | 2 | 2 | 3 | 0 | 0 |
| **Feedback part 2** | **Very unsure** | **Somewhat unsure** | **Neutral** | **Somewhat confident** | **Very confident** |  |
| **Confidence in completing survey questions** | 3 | 5 | 3 | 4 | 0 |  |

Feedback part 1 "high use": for training material use, corresponds to use of material to answer most questions; for external research use, corresponds to 1+ hours of independent research. Feedback part 2 response options: Very unsure – I struggled to answer the majority of the questions; Somewhat unsure – I had difficulty answering several questions; Neutral – I didn't feel particularly confident or unsure; Somewhat confident – I was fairly certain, but unsure about a few questions; Very confident – I felt sure about the majority of my answers

### Sensitivity analyses

We conducted post-hoc sensitivity analyses to assess potential drivers of the differences in effect size estimate between experts. Removing outliers, defined as best-guess responses outside 1.5 times the IQR (on the log-scale) for each question individually, reduced median effect size estimates by up to 50%, compared to the main analysis (Supplementary Table 6). Excluding experts who were “very unsure” about survey responses generally produced larger pooled estimates than the main analysis, for quitting and switching questions (Supplementary Table 7). Stratifying by high versus low use of training material (per Table 4) did not produce a consistent direction of change compared to the main analysis (Supplementary Table 8).

## Discussion

In this study, we applied structured EKE to quantify plausible behavioural responses to a VLNC policy in Australia, including cessation, switching to vaping or dual use, smoking uptake and illicit tobacco sourcing. Across 15 experts, pooled responses suggested that a VLNC mandate would increase smoking cessation and switching to e-cigarette use, with larger effects in the first year than in subsequent years of the policy. Estimates for smoking and vaping uptake were more variable. Experts also estimated substantial switching to illicit sourcing of normal-nicotine cigarettes, highlighting strong illicit tobacco control as a prerequisite prior to and after the introduction of a VLNC policy to ensure policy effectiveness.

Responses were collected separately with no interaction between experts then mathematically pooled and should therefore not be interpreted as consensus (as is generally achieved through behavioural aggregation, or a Delphi process). There was large variation in responses for most questions, with greater variation for the questions on switching from smoking to dual use, and uptake of illicit tobacco (see Supplementary Table 9 for 2.5^th^ and 97.5^th^ percentiles of pooled distributions). Further, while for smoking cessation questions the direction of effect was the same across experts (an increase under the intervention relative to BAU), this was not the case for dual use cessation and e-cigarette switching, with some experts including a potential decrease in these behaviours in their estimates. The same was seen for the smoking uptake, with some experts including an increase under the policy in their estimates. The latter may have reflected an assumed increase in illicit market uptake.

A clear divergence in responses emerged between experts of the Australian tobacco policy context and other country experts. Australian experts estimated smaller relative impacts on quitting and switching, no reduction of smoking uptake, and much higher transitions to illicit sourcing under the VLNC policy. This likely reflects context specific knowledge, particularly the recent growth in illicit tobacco use. The most recent estimate that illicit tobacco accounts for 50-60% of total tobacco consumption in Australia [14] was not available when the survey and training material were prepared – experts were provided with earlier estimates from the Australian Taxation Office, placing illicit market share at around 18% of total tobacco use.[25] Non-Australian experts may therefore have underestimated the current scale of Australia’s illicit market. However, it is also possible that Australian experts were less familiar with the VLNC proposal, as it has not been a prominent policy in Australian tobacco control debates compared to settings such as the US and New Zealand. Therefore, experts from Australia may have underestimated the impact of a VLNC policy compared to international experts.

The findings of this analysis will inform simulation modelling of a denicotinisation policy in Australia using the SHINE-Tobacco modelling platform.[20,21] The model will incorporate the wide uncertainty in responses across experts and include scenario analyses, for example estimating expected policy impacts based on the more conservative Australian sub-sample responses, and comparing results to intervention parameterisation with other sources of VLNC evidence (e.g. RCTs, surveys among people who smoke). Further, the model can be used to explore competing behavioural pathways. For example, experts estimated larger increases in switching from smoking to dual use than quitting all nicotine or switching to exclusive vaping. If people compensate for lower nicotine exposure by adding e-cigarette use while continuing to smoke VLNC cigarettes, the net impact on smoking prevalence and health gains may be smaller than cessation estimates alone imply. Simulation modelling can quantify the outcome of these competing effects. Finally, modelling can incorporate impacts on smoking intensity, which was not considered in this exercise.

These estimates can help compare denicotinisation with other endgame policies, particularly by distinguishing the likely impacts of policies that primarily affect smoking uptake from those that affect smoking cessation. This distinction is important for policymakers because impacts on prevalence, morbidity and mortality occur over different time horizons depending on the behavioural pathway and age group affected.

Alongside modelling, and in lieu of empirical evidence from a jurisdiction implementing a VLNC policy, future work may involve conducting similar expert elicitation exercises with law enforcement officials (given the clear illicit market link). Analyses including people who smoke are also needed, for example in the form of surveys or experimental marketplace analyses to investigate intended actions would be under a VLNC policy. Similar work has been conducted in settings outside of Australia [26,27]; however, as highlighted in the present analysis, context-specific information is needed.

A previous EKE on a US denicotinisation policy was published in 2018.[10] Pooled values from eight experts estimated that in the initial year of the policy, 19% (IQR 11-30%) of females and 21% (IQR 12-30%) of males would quit smoking (combining both quitting all nicotine and switching to non-combustible products), with estimates of 13.5% (8.4-23.8%) and 15% (9.4-26.3%) in subsequent years. A greater proportion of these total estimates were for quitting all nicotine than switching to non-combustible products [10], in opposition to our findings. However, like our study, the US experts anticipated larger shifts to dual use compared to quitting all nicotine. The US study experts also estimated a larger decrease in smoking uptake under the policy, with a median 50% reduction in both current and subsequent years (compared to BAU), and a median 37.5% of those deterred from smoking taking up use of a non-combustible product instead.[10] Importantly, the US study explicitly instructed participants to assume there was no illicit supply of normal-nicotine tobacco under the policy, which may account for some of the difference to our findings.[10]

Experts who were more supportive of novel endgame policies, or specifically a VLNC policy, may have given larger effect size estimates for the policy. However, excluding participants who did not respond that endgame policies were needed to achieve a tobacco endgame in their country generally resulted in more conservative pooled estimates. This suggests that support for endgame policy did not inflate expectations of this policy’s potential effect, although the survey did not measure support for a VLNC policy specifically.

Several limitations to our study should be considered. First, estimating the potential population impact of a nationwide denicotinisation policy is inherently difficult. While the survey was first piloted among the study investigators, participants still reported difficulty in completing the survey. Nine of the 15 experts who completed the survey reported being very or somewhat unsure about their responses. Further, some experts provided feedback that they would have preferred to select an “I don’t know” response option for some questions, but this was not available. One invited expert declined to complete the survey due to concerns around speculating policy impacts.

The results for the smoking and vaping uptake questions should be interpreted with caution. Some experts (n=3) gave non-zero values for vaping uptake substitution estimates despite previously estimating no reduction, or an increase, in smoking uptake (in those cases, there would be no group deterred from smoking to substitute with vaping). There was also evidence of potential confusion surrounding the illicit market questions. Values up to and higher than 90% switching to illicit market cigarettes with normal nicotine content were provided as responses. The question was intended to provide estimates of the percentage who would switch to illicit cigarettes among all people who smoked prior to the intervention. However, the very high percentages in some responses suggest that some experts interpreted this question as the proportion switching to illicit cigarettes among those who did not quit smoking or switch to vaping in response to policy implementation.

The limitation of EKE as a source of evidence should also be acknowledged. We cannot assume that the pooled median responses reflect the true policy impact. What this approach can do is provide an estimate of policy impact and characterise the range of uncertainty in opinion on the policy from appropriate experts and determine where major risks lie. Improved estimates of the impact of a mandated nicotine reduction will likely require evidence from implementation of the policy. However, waiting for such real-world evidence to be generated could mean policy-paralysis for an option that may have the potential for achieving large health gains. Hence, alternative evidence such as that gathered in this analysis is warranted.[28] These findings, and future related modelling, can help to build on existing evidence and support an assessment of the risks and benefits of implementing this policy in Australia.

## Conclusion

Given the limited ability of existing short-duration VLNC trials to predict population-level effects of a mandatory product standard, and the absence of real-world implementation evidence, this study provides Australia-specific estimates of the potential behavioural effects of a VLNC policy. The EKE estimates suggest that denicotinisation could accelerate progress towards a tobacco endgame, but that its benefits may be constrained by Australia’s policy environment, particularly illicit access to nicotine-containing cigarettes. Our pooled estimates should not be interpreted as a consensus forecast or treated as being equivalent to empirical data. Rather, the value of the EKE approach is that it defines a plausible range of behavioural responses that simulation models can test, to help identify optimal policy choices. The findings of this paper indicate that denicotinisation warrants consideration, provided implementation is accompanied by robust illicit-market control and monitoring of substitution pathways. The evidence from this analysis should be updated with real-world, empirical evidence as it becomes available.

## Data Availability

Aggregated data produced in the present work are contained in the manuscript. Individual level data is not available, to maintain expert participant confidentiality and in line with the study protocol.

## Acknowledgements

We thank all experts who participated in the EKE for generously contribution their time and expertise to this study. Experts who consented to be acknowledged by name are listed in the Supplementary Results.

## Declaration of Interests

SH is supported by a Research Training Program Scholarship administered by the University of Melbourne and a PhD top-up scholarship from the NHMRC Centre of Research Excellence on Achieving the Tobacco Endgame (CREATE; GNT1198301). SH is the Communications Officer for the Society of Research on Nicotine and Tobacco (SRNT) Oceania Chapter (2025-present).

RE is the current president of the SRNT Oceania Chapter (2025-present).

TW receives a fellowship from the NHMRC CREATE (GNT1198301).

## Funding

This research was funded through an NHMRC grant (GNT1198301). The funders were not involved in any phase of the study.

## Data availability

Data not publicly available

## Notes

### Clinical Protocols

https://osf.io/mndte/overview

### Author Declarations

The Human Research Ethics Committee of the University of Melbourne gave ethical approval for this work (reference number: 2025-32045-69755-3).

