## Supplementary Material for "Estimating the impact of a denicotinisation policy on smoking behaviours in Australia – an expert knowledge elicitation study"

### Supplementary Methods

#### Literature review

At the outset of this project, a search of the literature was conducted to identify existing research on the effect of reduced nicotine content cigarettes. This information was used to guide formulation of the EKE survey and to provide the necessary background material for expert participants.

The research question of interest was: what is the effectiveness of low or very low nicotine content cigarettes compared to no intervention, or other smoking cessation interventions, on cigarette smoking behaviour (including at least one of number of cigarettes smoked, quit attempts made or successful cessation) among adults who currently smoke?

Given the novelty of the intervention, we kept the criteria for the search relatively broad.

**Inclusion criteria:**

- Population: adults (15+) who currently smoke cigarettes
- Intervention: low or very low nicotine content cigarettes
- Comparator: either no intervention, or a standard smoking cessation intervention (including nicotine replacement therapy, counselling, or use of e-cigarettes)
- Outcomes: at least one of number of cigarettes smoked, quit attempts made, or successful cessation AND rate of non-adherence to treatment or comparator, rate of study drop-out
- Study design: randomised controlled trials (RCTs) or systematic reviews of RCTs
- Time frame: minimum 30 days
- Years considered: no restriction
- Publication status: published and pre-print RCTs/systematic reviews
- Language: published in English or available to translate to English

**Exclusion criteria:**

- Population: specific sub-population of adults who currently smoke, for example those with mental illness, or during pregnancy. Exception: studies focusing on particular age groups, e.g., youth (15–24-year-olds)
- Comparator: no comparator/control group
- Outcomes: only reports subjective outcome (e.g., cigarette rating or preference, support for reduced nicotine policy)

The search was conducted on 21^st^ August 2024 (then updated on 17/03/26). The search terms, shown in Supplementary Table 1, were adapted from a systematic review by Klemperer et al. [1]

**Supplementary Table 1. Search terms by database.**

| **Database** | **Search Terms** | **Results** |
| --- | --- | --- |
| **Scopus** | TITLE-ABS-KEY ( smok* OR nicotin* OR tobacco OR cigarette* )  AND  TITLE-ABS-KEY ( "quit attempt" OR "harm reduction" OR quit* OR stop* OR ceas* OR cessation OR abstain OR abstinence OR discontinu* OR "give up" OR "smoking cessation agent" OR "smoking reduction" OR “**reduced exposure tobacco products” )**  AND  TITLE-ABS-KEY ( "very low nicotine" OR "reduced nicotine" OR "low nicotine" OR "denicotin*" ) | 533 |
| **PubMed** | **("very low nicotine"[Title/Abstract] OR "reduced nicotine"[Title/Abstract] OR "low nicotine"[Title/Abstract] OR "denicotinis*"[Title/Abstract] OR "denicotiniz*"[Title/Abstract] OR “reduced exposure tobacco products”)**  **AND**  **(cigarette smoking[MeSH Terms] OR tobacco dependence[MeSH Terms] OR tobacco[MeSH Terms] OR nicotine[MeSH Terms] OR tobacco use[MeSH Terms] OR smoking[MeSH Terms] OR smok* [Title/Abstract] OR nicotin* [Title/Abstract] OR tobacco [Title/Abstract]))**  **AND**  **("quit attempt" [Title/Abstract] OR "harm reduction" [Title/Abstract] OR quit* [Title/Abstract] OR stop* [Title/Abstract] OR ceas* [Title/Abstract] OR Cessation [Title/Abstract] OR Abstain [Title/Abstract] OR Abstinence [Title/Abstract] OR Smoking Cessation [MeSH] OR Tobacco Use Cessation [MeSH] OR Smoking Cessation Agents [MeSH] OR discontinu* [Title/Abstract] OR "give up" [Title/Abstract] OR smoking reduction[MeSH Terms])** | 478 |
| **Total (after removing duplicates)** | | **570** |

From the search, one systematic review was identified, specifically of people who aren’t ready/motivated to quit smoking. We identified 76 RCTs, with 36 of these being original publications and the rest being secondary analyses.

Most RCTs were conducted in the US. Two trials were identified in Aotearoa/New Zealand, one in England, and one in China. Many studies looked at specific at-risk populations, including those with serious mental illnesses, low socioeconomic groups, co-users of tobacco and cannabis, and youth. The main outcomes observed were smoking intensity (cigarettes smoked per day (CPD)), markers of withdrawal, dependence and cravings. Other outcomes include examination of compensatory behaviours, compliance with the treatment allocation, and product preferences. Few studies were large enough or continued for a long enough duration to assess quit rates or long-term abstinence between intervention and control arms.

The RCTs that met inclusion criteria are summarised in Table 1 of Participant Training Material A (provided as Supplementary Material). Note that individual studies included within the identified systematic review are not reported on separately.

#### Recruitment

Search terms used to identify potential expert participants are shown in Supplementary Table 2.

**Supplementary Table 2. Search terms for Scopus expert search**

| **Topic** | **Search terms*** |
| --- | --- |
| **VLNC research** | "very low nicotine" OR "low nicotine" OR "reduced nicotine" OR “denicotin*” |
| **Nicotine addiction** | “addiction” AND (“nicotine” OR “tobacco”) AND (“cessation” OR “quit”) |
| **Tobacco control policy** | (“tobacco control” OR “tobacco endgame”) AND “policy” |
| **Illicit tobacco/nicotine markets** | “illicit tobacco market” OR (“illicit” AND “tobacco”) OR (“illicit” AND “nicotine”) OR (“illicit AND e-cigarette*”) |

*Title-abstract-key word search

#### Survey details

The full list of survey questions are provided in a separate Supplementary Document.

Two examples of the survey, as presented in Qualtrics, are shown below. In example A (Supplementary Figure 1), experts are asked to provide the smoking cessation rate in the first year of the policy for 30+ year olds. The cessation rate ‘reference’ without the policy is provided (red ‘HELPER’ text below question). Note that the ‘best guess’ estimate (Q1c) question box does not appear until both the minimum and maximum have been provided. A pop-up box (green text below response) shows what the best guess estimate means in relative terms.

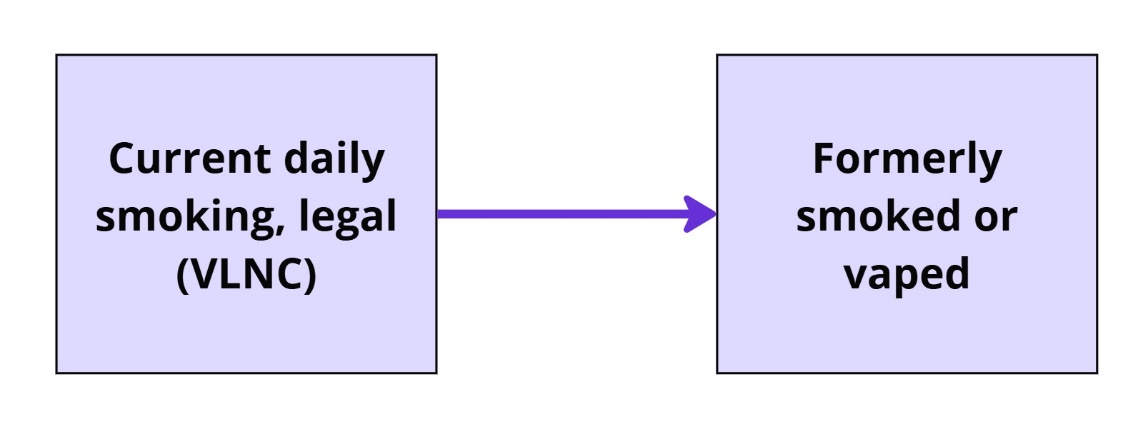

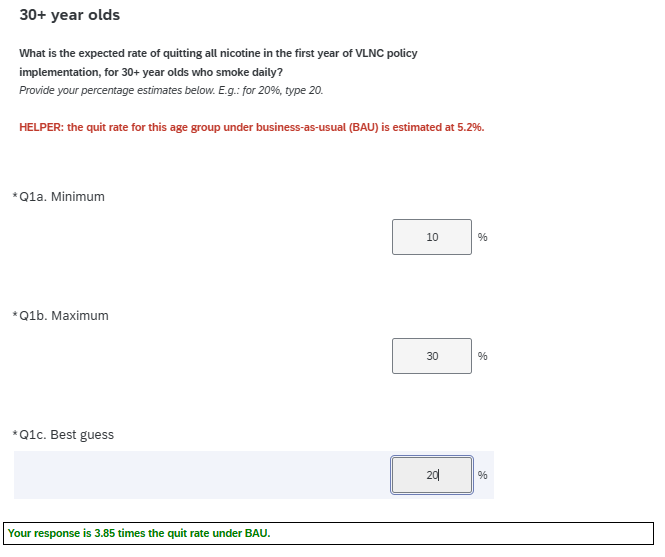

**Supplementary Figure 1. Qualtrics survey question and response: Example A**

The reference values provided throughout the survey are supplied in Supplementary Table 3. These values were derived from the SHINE Tobacco simulation model, which estimated smoking uptake, quit, and switch rates based on smoking and vaping prevalence by age and over time. [2]

**Supplementary Table 3. Reference values for base year (2023)**

| **Transition** | **<30-year-olds** | **30+ year olds** |
| --- | --- | --- |
| **Smoking (S)** $\boldsymbol{\to}$ **Former Smoking (FS)** | 2.1% | 5.2% |
| **Smoking (S)** $\boldsymbol{\to}$ **Vaping (V)** | 2.9% | 2.1% |
| **No product use** $\boldsymbol{\to}$ **Smoking (S)** | 1.0% | NA |

In Example B (Supplementary Figure 2), experts are asked to estimate the relative impact of the policy, in the first year for 30+ year olds, on switching from smoking to dual use. Rather than using a pre-determined reference point, as for the previous example, the response from a previous question within the survey is provided as a reference (red text below question). Once the best guess has been provided, a pop-up box (green text below response) shows what the response means in relation to the previous question.

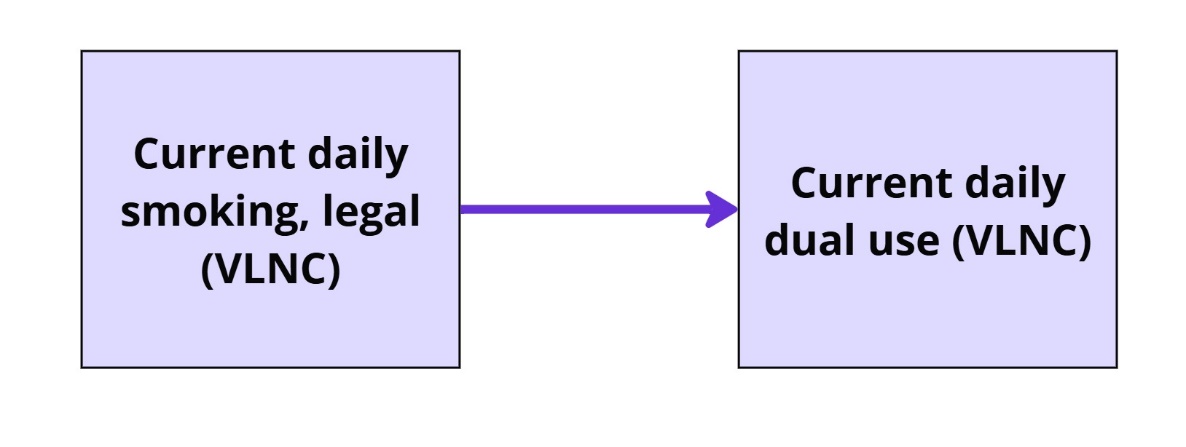

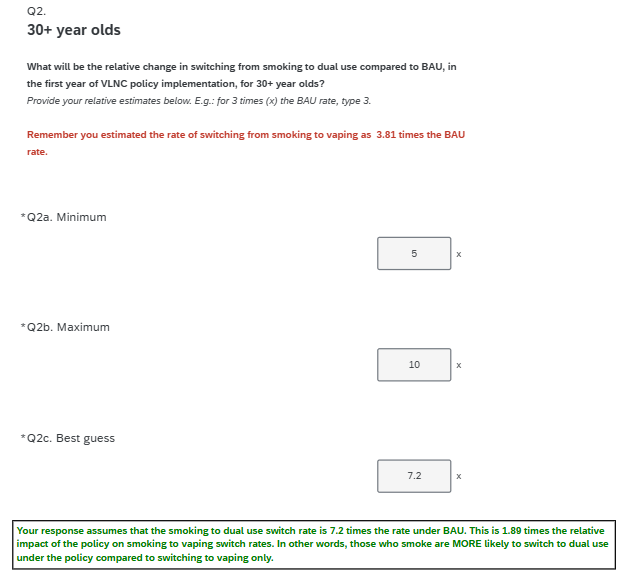

**Supplementary Figure 2. Qualtrics survey question and response: Example B.**

### Supplementary Results

#### Participating experts

Please note that the below named experts are those who consented to be acknowledged in this paper. We thank all participating experts for their contribution to this work.

[Note for reviewers and editors: the names of experts have been removed from this document for the peer review process].

#### Quantitative research expertise

**Supplementary Table 4. Pooled results: median impact (and interquartile range) on smoking and vaping behaviours for experts reporting quantitative research expertise (n=12).**

| **Transition** | **<30-year-olds** | | **30+ year olds** | |
| --- | --- | --- | --- | --- |
|  | **First year of policy** | **Subsequent years** | **First year of policy** | **Subsequent years** |
| **Relative change compared to business-as-usual** | | | | |
| **Smoking** $\boldsymbol{\to}$  **Former Smoking** | 3.8 (2.2-7.1) | 3.6 (2-5.7) | 2.1 (1.3-4) | 2.5 (1.3-3.8) |
| **Dual use** $\boldsymbol{\to}$  **Former Smoking** | 2.1 (1.1-3.7) | 2.4 (1.1-4.4) | 1.6 (1.1-4.1) | 1.4 (1-3.9) |
| **Smoking** $\boldsymbol{\to}$  **Vaping** | 4.6 (2.2-14.9) | 3.8 (1.4-14.7) | 4.4 (2.7-10.2) | 3.6 (1.6-10.1) |
| **Smoking** $\boldsymbol{\to}$  **Dual use** | 4.6 (2.4-29.7) | 4.2 (2.1-27.5) | 4.2 (2.7-26.9) | 2.9 (1.8-25.2) |
| **No product use** $\boldsymbol{\to}$  **Smoking*** | 0.7 (0.1-1) | 0.7 (0.3-0.8) | N/A | N/A |
| **Percentage change compared to business-as-usual** | | | | |
| **Legal smoking** $\boldsymbol{\to}$  **Illicit market^#^** | 35.8% (15%-53.1%) | N/A | 32.1% (14.1%-46%) | N/A |
| **Legal dual use** $\boldsymbol{\to}$  **Illicit market^#^** | 20.2% (13.1%-57.8%) | N/A | 20.1% (6.9%-42.1%) | N/A |

*Relative impact on smoking uptake only considered for <30-year-olds (ages for which majority of uptake occurs).

^#^ Note that, while in Australia e-cigarettes are banned from sale in retail settings (except for pharmacy access for smoking cessation), in this study the illicit market refers to the tobacco illicit market specifically.

Relative risk values <1 indicate lower transition rates under the intervention compared to BAU, while values >1 indicate a higher transition rate under the intervention compared to BAU. Interquartile range estimated by linear pooling of individual response distributions with the SHELF package in R; individual responses assumed to follow log-normal distributions, where best guess = 50^th^ percentile, and the minimum and maximum responses are the 2.5^th^ and 97.5^th^ percentiles.

#### Tobacco endgame support

**Supplementary Table 5. Pooled results: median impact (and interquartile range) on smoking and vaping behaviours for experts indicating support for novel tobacco control policies (n=12)**

| **Transition** | **<30-year-olds** | | **30+ year olds** | |
| --- | --- | --- | --- | --- |
|  | **First year of policy** | **Subsequent years** | **First year of policy** | **Subsequent years** |
| **Relative change compared to business-as-usual** | | | | |
| **Smoking** $\boldsymbol{\to}$  **Former Smoking** | 3.5 (1.2-9.8) | 2.4 (1-5.9) | 2 (1.2-5.3) | 1.9 (1.1-3.8) |
| **Dual use** $\boldsymbol{\to}$  **Former Smoking** | 1.9 (1.1-3.6) | 2.3 (1-4.2) | 2 (1.1-4.1) | 1.5 (1-3.6) |
| **Smoking** $\boldsymbol{\to}$  **Vaping** | 2.7 (1.2-13.2) | 2 (1.1-8.1) | 3.3 (1.3-8.3) | 1.7 (1.1-6) |
| **Smoking** $\boldsymbol{\to}$  **Dual use** | 3 (2-18.5) | 2.5 (1-16.3) | 3.3 (1.7-14.3) | 1.9 (1-9.6) |
| **No product use** $\boldsymbol{\to}$  **Smoking*** | 0.8 (0.1-1) | 0.7 (0.1-0.8) | N/A | N/A |
| **Percentage change compared to business-as-usual** | | | | |
| **Legal smoking** $\boldsymbol{\to}$  **Illicit market^#^** | 41.2% (13.2%-53.2%) | N/A | 38.2% (21.1%-51.6%) | N/A |
| **Legal dual use** $\boldsymbol{\to}$  **Illicit market^#^** | 30% (14.3%-54.4%) | N/A | 23.9% (10.7%-42.2%) | N/A |

*Relative impact on smoking uptake only considered for <30-year-olds (ages for which majority of uptake occurs).

^#^ Note that while in Australia, e-cigarettes are banned from sale in retail settings (except for pharmacy access for smoking cessation), In this study the illicit market refers to the tobacco illicit market specifically.

Relative risk values <1 indicate lower transition rates under the intervention compared to BAU, while values >1 indicate a higher transition rate under the intervention compared to BAU. Interquartile range estimated by linear pooling of individual response distributions with the SHELF package in R; individual responses assumed to follow log-normal distributions, where best guess = 50^th^ percentile, and the minimum and maximum responses are the 2.5^th^ and 97.5^th^ percentiles.

#### Outliers removed

**Supplementary Table 6. Pooled results: median impact (and interquartile range) on smoking and vaping behaviours for, with outliers^#^ for each question removed**

| **Transition** | **<30-year-olds** | | **30+ year olds** | |
| --- | --- | --- | --- | --- |
|  | **First year of policy** | **Subsequent years** | **First year of policy** | **Subsequent years** |
| **Relative change compared to business-as-usual** | | | | |
| **Smoking** $\boldsymbol{\to}$  **Former Smoking** | 3 (1.3-4) | 2.4 (1.1-3.8) | 1.7 (1.2-2.9) | 1.5 (1-2.9) |
| **Dual use** $\boldsymbol{\to}$  **Former Smoking** | 1.9 (1.1-2.6) | 1.9 (1-2.8) | 1.4 (1-2.8) | 1.3 (1-2.6) |
| **Smoking** $\boldsymbol{\to}$  **Vaping** | 2.5 (1.2-4.9) | 1.4 (1-3.5) | 3 (1.3-4.6) | 1.5 (1.1-3.3) |
| **Smoking** $\boldsymbol{\to}$  **Dual use** | 3.4 (2.1-7.1) | 2.8 (1.2-24.3) | 3 (1.6-4.6) | 1.7 (1-2.7) |
| **No product use** $\boldsymbol{\to}$  **Smoking*** | 0.7 (0-1) | 0.7 (0.5-1) | N/A | N/A |
| **Percentage change compared to business-as-usual** | | | | |
| **Legal smoking** $\boldsymbol{\to}$  **Illicit market^** | 20.8% (14.1%-47.1%) | N/A | 32.1% (15.5%-46.4%) | N/A |
| **Legal dual use** $\boldsymbol{\to}$  **Illicit market^** | 20.3% (13.7%-45.8%) | N/A | 20.1% (7.9%-34.6%) | N/A |

*Relative impact on smoking uptake only considered for <30-year-olds (ages for which majority of uptake occurs).

^#^Outliers defined as values outside 1.5 times the interquartile range (IQR).

^Note that, while in Australia e-cigarettes are banned from sale in retail settings (except for pharmacy access for smoking cessation), in this study the illicit market refers to the tobacco illicit market specifically.

Relative risk values <1 indicate lower transition rates under the intervention compared to BAU, while values >1 indicate a higher transition rate under the intervention compared to BAU. Interquartile range estimated by linear pooling of individual response distributions with the SHELF package in R; individual responses assumed to follow log-normal distributions, where best guess = 50^th^ percentile, and the minimum and maximum responses are the 2.5^th^ and 97.5^th^ percentiles.

#### ‘Unsure’ responses removed

**Supplementary Table 7. Pooled results: median impact (and interquartile range) on smoking and vaping behaviours for experts who did not report being very unsure about survey questions (n=12)**

| **Transition** | **<30-year-olds** | | **30+ year olds** | |
| --- | --- | --- | --- | --- |
|  | **First year of policy** | **Subsequent years** | **First year of policy** | **Subsequent years** |
| **Relative change compared to business-as-usual** | | | | |
| **Smoking** $\boldsymbol{\to}$  **Former Smoking** | 3.8 (1.3-8.4) | 3.4 (1.1-5.4) | 2.7 (1.4-5.3) | 1.9 (1.1-3.8) |
| **Dual use** $\boldsymbol{\to}$  **Former Smoking** | 1.9 (1.1-3.6) | 1.9 (1-4.2) | 1.4 (1-3.2) | 1.4 (1-3.9) |
| **Smoking** $\boldsymbol{\to}$  **Vaping** | 4.7 (1.8-13.2) | 3 (1.1-8) | 4.5 (2-8.3) | 3.1 (1.1-6.3) |
| **Smoking** $\boldsymbol{\to}$  **Dual use** | 4.7 (2.6-27.7) | 3 (1-26.5) | 4.4 (2.4-14.3) | 2.5 (1-11) |
| **No product use** $\boldsymbol{\to}$  **Smoking*** | 0.7 (0-1) | 0.5 (0.1-0.8) | N/A | N/A |
| **Percentage change compared to business-as-usual** | | | | |
| **Legal smoking** $\boldsymbol{\to}$  **Illicit market^#^** | 18.7% (13.2%-50.2%) | N/A | 33.6% (14.6%-51.4%) | N/A |
| **Legal dual use** $\boldsymbol{\to}$  **Illicit market^#^** | 23.9% (13.5%-55%) | N/A | 20.7% (6.9%-37.7%) | N/A |

*Relative impact on smoking uptake only considered for <30-year-olds (ages for which majority of uptake occurs).

^#^ Note that, while in Australia e-cigarettes are banned from sale in retail settings (except for pharmacy access for smoking cessation), in this study the illicit market refers to the tobacco illicit market specifically.

Relative risk values <1 indicate lower transition rates under the intervention compared to BAU, while values >1 indicate a higher transition rate under the intervention compared to BAU. Interquartile range estimated by linear pooling of individual response distributions with the SHELF package in R; individual responses assumed to follow log-normal distributions, where best guess = 50^th^ percentile, and the minimum and maximum responses are the 2.5^th^ and 97.5^th^ percentiles.

#### Training material use

**Supplementary Table 8. Pooled results: median impact (and interquartile range) on smoking and vaping behaviours for experts who reported moderate-high vs. low training material use**

| **Transition** | **<30-year-olds** | | **30+ year olds** | |
| --- | --- | --- | --- | --- |
|  | **First year of policy** | **Subsequent years** | **First year of policy** | **Subsequent years** |
| **Moderate-high use of training material (n=6)** | | | | |
| **Relative change compared to business-as-usual** | | | | |
| **Smoking** $\boldsymbol{\to}$  **Former Smoking** | 2.4 (1.7-4.1) | 2.9 (1.8-4.1) | 1.8 (1.4-2.3) | 1.8 (1.1-2.8) |
| **Dual use** $\boldsymbol{\to}$  **Former Smoking** | 1.9 (1.4-2.6) | 2.1 (1-2.8) | 1.2 (1-2) | 1.3 (1-2.5) |
| **Smoking** $\boldsymbol{\to}$  **Vaping** | 4.6 (1.7-9.5) | 3 (1.2-5.1) | 4.3 (2.4-6.1) | 2 (1.2-5.1) |
| **Smoking** $\boldsymbol{\to}$  **Dual use** | 4 (2.5-26.7) | 4.2 (2.7-25.7) | 2.9 (1.8-6.4) | 2.3 (1.7-6.4) |
| **No product use** $\boldsymbol{\to}$  **Smoking*** | 0.8 (0.4-1) | 0.7 (0.7-0.8) | N/A | N/A |
| **Percentage change compared to business-as-usual** | | | | |
| **Legal smoking** $\boldsymbol{\to}$  **Illicit market^#^** | 18.4% (12.2%-42.8%) | N/A | 27.4% (19.8%-37.5%) | N/A |
| **Legal dual use** $\boldsymbol{\to}$  **Illicit market^#^** | 17.3% (13.1%-44.2%) | N/A | 24.1% (17.6%-34.6%) | N/A |
| **Low use of training material (n=9)** | | | | |
| **Relative change compared to business-as-usual** | | | | |
| **Smoking** $\boldsymbol{\to}$  **Former Smoking** | 3.8 (1.1-7.7) | 2.9 (1.1-5.8) | 2.9 (1.1-5) | 1.8 (1.1-3.3) |
| **Dual use** $\boldsymbol{\to}$  **Former Smoking** | 1.9 (1-4.7) | 1.9 (1-5.2) | 2 (1.1-6) | 1.4 (1-4.3) |
| **Smoking** $\boldsymbol{\to}$  **Vaping** | 2.9 (1.2-6.6) | 2.7 (1.1-5.2) | 3.5 (1.4-5.5) | 2.8 (1.1-4.8) |
| **Smoking** $\boldsymbol{\to}$  **Dual use** | 3.8 (1.8-6.6) | 2 (1-5.2) | 4 (2.5-5.6) | 2 (1-4.8) |
| **No product use** $\boldsymbol{\to}$  **Smoking*** | 0.5 (0-1) | 0.5 (0.1-0.8) | N/A | N/A |
| **Percentage change compared to business-as-usual** | | | | |
| **Legal smoking** $\boldsymbol{\to}$  **Illicit market^#^** | 39.9% (15%-56.5%) | N/A | 40.1% (15%-51.2%) | N/A |
| **Legal dual use** $\boldsymbol{\to}$  **Illicit market^#^** | 30% (16.3%-76.7%) | N/A | 16.6% (7.3%-44.5%) | N/A |

*Relative impact on smoking uptake only considered for <30-year-olds (ages for which majority of uptake occurs).

^#^ Note that, while in Australia e-cigarettes are banned from sale in retail settings (except for pharmacy access for smoking cessation), In this study the illicit market refers to the tobacco illicit market specifically.

Relative risk values <1 indicate lower transition rates under the intervention compared to BAU, while values >1 indicate a higher transition rate under the intervention compared to BAU. Interquartile range estimated by linear pooling of individual response distributions with the SHELF package in R; individual responses assumed to follow log-normal distributions, where best guess = 50^th^ percentile, and the minimum and maximum responses are the 2.5^th^ and 97.5^th^ percentiles.

#### 2.5^th^-97.5^th^ percentile results

**Supplementary Table 9. Pooled results: 2.5^th^ and 97.5^th^ percentile responses on smoking and vaping behaviours (n=16)**

| **Transition** | **<30-year-olds** | | **30+ year olds** | |
| --- | --- | --- | --- | --- |
|  | **First year of policy** | **Subsequent years** | **First year of policy** | **Subsequent years** |
| **Relative change compared to business-as-usual** | | | | |
| **Smoking** $\boldsymbol{\to}$  **Former Smoking** | 1-27.8 | 1-12.6 | 1-12.9 | 1-8.3 |
| **Dual use** $\boldsymbol{\to}$  **Former Smoking** | 0.9-11.7 | 0.4-11.7 | 0.8-21.4 | 0.8-21.7 |
| **Smoking** $\boldsymbol{\to}$  **Vaping** | 0.4-27.6 | 0.4-27.6 | 1.1-24.8 | 0.9-21 |
| **Smoking** $\boldsymbol{\to}$  **Dual use** | 1.2-77.9 | 1.0-41.6 | 1.1-63.7 | 1-42.2 |
| **No product use** $\boldsymbol{\to}$  **Smoking*** | <0.01-1.7 | 0.1-1.2 | N/A | N/A |
| **Percentage change compared to business-as-usual** | | | | |
| **Legal smoking** $\boldsymbol{\to}$  **Illicit market^#^** | 4.4%-95.8% | N/A | 8.6%-95.8% | N/A |
| **Legal dual use** $\boldsymbol{\to}$  **Illicit market^#^** | 3.8%-96.2% | N/A | 3.2%-95.8% | N/A |

*Relative impact on smoking uptake only considered for <30-year-olds (ages for which majority of uptake occurs).

^#^ Note that while in Australia, e-cigarettes are banned from sale in retail settings (except for pharmacy access for smoking cessation), In this study the illicit market refers to the tobacco illicit market specifically.

Relative risk values <1 indicate lower transition rates under the intervention compared to BAU, while values >1 indicate a higher transition rate under the intervention compared to BAU. 2.5^th^ and 97.5^th^ percentile values estimated by linear pooling of individual response distributions with the SHELF package in R; individual responses assumed to follow log-normal distributions, where best guess = 50^th^ percentile, and the minimum and maximum responses are the 2.5^th^ and 97.5^th^ percentiles.
